# Did the COVID-19 pandemic result in more family physicians stopping practice? Results from Ontario, Canada

**DOI:** 10.1101/2021.09.21.21263891

**Authors:** Tara Kiran, Michael E. Green, Fangyun C. Wu, Alexander Kopp, Lidija Latifovic, Eliot Frymire, Richard H. Glazier

**Affiliations:** Department of Family and Community Medicine, St. Michael’s Hospital, University of Toronto, Toronto, Ontario; MAP Centre for Urban Health Solutions, St. Michael’s Hospital, Toronto, Ontario; ICES Central, Toronto, Ontario; Institute of Health Policy, Management and Evaluation, University of Toronto, Toronto, Ontario; Dept of Family Medicine, Queen’s University, Kingston, ON; Health Services and Policy Research Institute, Queens University, Kingston, ON; ICES Queens, Kingston, Ontario; Dalla Lana School of Public Health, University of Toronto, Toronto, ON

## Abstract

**Purpose:** To understand changes in family physician practice patterns and whether more family physicians stopped working during the COVID-19 pandemic compared to previous years.

**Methods:** We analyzed administrative data from Ontario, Canada two ways: cross-sectional and longitudinal. First, we identified the percentage and characteristics of all family physicians who had a minimum of 50 billing days in 2019 but no billings during the first six months of the pandemic. Second, for each year from 2010 to 2020, we calculated the percentage of physicians who billed for services in the first quarter of the calendar year but submitted no bills between April and September of the given year.

**Results:** We found 3.1% of physicians working in 2019 (N=385/12,247) reported no billings in the first six months of the pandemic. Compared with other family physicians, a higher portion were age 75 or older (13.0% vs. 3.4%, p<0.001), had fee-for-service reimbursement (38% vs 25%, p<0.001), and had a panel size under 500 patients (40% vs 25%, p<0.001). Between 2010 and 2019, an average of 1.6% of physicians who practiced in the first quarter had no billings in each of the second and third quarters of the calendar year compared to 3.0% in 2020 (p<0.001).

**Conclusions:** Approximately twice as many family physicians stopped work in Ontario, Canada during COVID-19 compared to previous years, but the absolute number was small and those who did had smaller patient panels. More research is needed to understand the impact on primary care attachment and access to care.

## Introduction

The COVID-19 pandemic disrupted how care was delivered by family physicians. To keep patients and staff safe during in-person visits, practices adopted a range of infection and control measures including active and passive screening, environmental cleaning, strict use of personal protective equipment, and reducing the number of staff and patients in the office. There was a dramatic shift to virtual care which increased 56-fold and early on comprised approximately 70% of total visits.^1^ In the first few weeks, when healthcare systems were worried about hospitals being overwhelmed by patients with COVID-19, non-essential care was deferred^2^ and total visits to primary care decreased by nearly 30%.^1^ At the same time, primary care physicians were tasked with other system roles including staffing of COVID-19 assessment centres.^3^

These changes placed extraordinarily stress on family physicians, particularly in Canada and the US, where many operate as small business owners and rely on fee-for-service billings for revenue.^4^ Surveys of family physicians in the US and Canada during the first wave of the pandemic detail numerous challenges including reduced revenue, retention of office staff, and difficulty obtaining personal protective equipment; many also worried about their own personal safety.^5,6^

There have been concerns that these practice challenges have led some family physicians to prematurely stop working.^7^ In one US survey, almost twenty percent of family physicians reported having colleagues who retired early or were planning on it.^8^ In some Canadian jurisdictions, the regulatory colleges have noted complaints from patients about difficulties getting an appointment, especially in-person.^9^ Primary care is the front door to the healthcare system and a reduction in the workforce would have repercussions for population health.^10^ However, there has been little research to understand the extent to which the COVID-19 pandemic has resulted in family physicians stopping work, the variation in practice patterns, and related factors. Our study sought to understand variation in practice patterns during COVID-19 and the number and characteristics of physicians stopping work in the first six months of the pandemic.

## Methods

### Context and Settings

Ontario is Canada’s largest province with an estimated population of 14.7 million in 2021.^11^ Medically necessary physician and hospital visits are fully insured and free at the point-of-care for all permanent residents via the Ontario Health Insurance Plan. Primary care is largely delivered by family physicians, roughly 80% of whom work in patient enrolment models (PEMs).^12^ Physicians in PEMs formally enrol patients and largely work in groups of 3 or more physicians with shared after-hours responsibility. The three main types of PEMs differ by the amount of fee-for-service payment and funding for non-physician health professionals.^13^ Approximately 20% of primary care physicians do not belong to a PEM and operate independent, fee-for-service practices with many working in walk-in clinics or doing a focused practice (e.g. sports medicine).^14^

Shortly after the global pandemic was declared on March 11, 2020, the Ontario government issued guidance to the health sector to halt all non-essential health services and reduce in-person visits by transitioning to virtual care where possible. On March 14, the Ontario government introduced new virtual billing codes to compensate physicians for phone and video visits.^15^ These codes were in-basket for physicians paid largely through capitation. By early June, the health sector was asked to initiate a gradual resumption to full scope of services while still maximizing virtual care. Through the pandemic, family physicians were responsible for securing their own personal protective equipment, hand sanitizer, plexiglass, environmental cleaning solutions, and other products required for office infection prevention and control (IPAC). About three months into the pandemic, some of these supplies were available free of charge through a central government supported procurement portal. No additional financial support was provided to assist with implementing IPAC recommendations.

### Study design and population

We conducted two analyses using linked administrative data. First, we examined variation in practice patterns during the first six months of the COVID-19 pandemic (March 11^th^ to September 29^th^, 2020) compared with the same period in 2019. We included all family physicians or general practitioners who had at least 50 billing days in 2019. Second, we conducted a repeated cross-sectional analysis to understand how many physicians stopped working between April 1 to September 30 of each year between 2010 and 2020. For this longitudinal analysis, we included all family physicians or general practitioners who billed for service between January 1 to March 31 of the given year.

Datasets only included de-identified data that were linked using unique encoded identifiers and analyzed at ICES. ICES is an independent, non-profit research institute whose legal status under Ontario’s health information privacy law allows it to collect and analyze health care and demographic data, without consent, for health system evaluation and improvement.

### Definitions and data sources

We used the Ontario Health Insurance Plan (OHIP) database to extract physician claims data, the Corporate Provider Database (CPDB) for physician age, sex, and practice postal code, and the Primary Care Population (PCPOP) database for physician group, model of care, and panel size. Physicians in a Patient Enrolment Model (PEM) were nested within groups that were in turn nested within a practice type. For panel size, patients were attributed to physicians based on enrolment data; patients who were not enrolled were attributed to a physician using virtual rostering according to the highest billings for that patient (see Appendix 1). We used practice postal code to classify physicians by rurality using the Rurality Index of Ontario (0=big cities; 1-9=small cities; 10-39=small communities; 40 or more=rural areas). We included physician visits with the location listed as office, home, or phone.

### Analysis

First, we examined total visits for each week from January 1, 2020 to Sep 29, 2020 and the same time period in 2019, stratified by type of visit (office, home, virtual). For each physician, we calculated the ratio of total visits from March 11 to September 29, 2020 to total visits in the same period in 2019. For physicians practicing in a Patient Enrolment Model (PEM) with a total group size of 3 physicians or more, we assessed the variation in the ratio both within a group and between groups in the same practice type. We calculated an intraclass correlation coefficient from a three-level, intercept-only mixed linear model to understand how much of the total variance in the ratio of visits during the pandemic to the same period in 2019 was attributable to physician group and practice type. We identified physicians who had zero visits from March 11 to September 29, 2020 and compared their characteristics to those who had any visits during that time period using a t-test for mean, a Kruskal-Wallis test for median, and a Chi-squared test for other categorical variables. We mapped the percentage of family physicians with zero visits between March 11 to September 29, 2020 by sub-region.

Second, for each year from 2010 to 2020, we examined the cohort of physicians practicing in the first 3 quarters of each year and noted the number and percent who stopped practicing (i.e., had no billings) between April 1 and September 30. We used a chi-squared test for independence and treated time as the categorical variable to test the null hypothesis that the proportion of physicians stopping work was not dependent on the year. Analysis was done in SAS^®^ Enterprise Guide and graphs were produced in R v4.0.5.

## Results

We analyzed data for 12,247 family physicians practicing in Ontario in 2019 (Table 1). Their mean age was 51; 48% were female, 49% practiced in an urban area, they worked in a range of practice models, and the average panel size was 1096 patients. Figure 1 shows that total visits dropped precipitously in mid-March, 2020 but largely recovered to previous levels by the end of September. During the week of September 29^th^, there were 916,831 total visits in 2019, of which 97% were in-office, compared to 903,402 total visits in 2020, of which 40% were in-office.

**Table 1.** Characteristics of all family physicians active in 2019 and comparison of characteristics between those with no outpatient visits during the first six months of the COVID-19 pandemic (March - September 2020) and those with any visits.

| Characteristic | All family physicians active in 2019 | Family physicians with any outpatient visits from March to September 2020 | Family physicians with no outpatient visits March to September 2020 | p-value |
| --- | --- | --- | --- | --- |
|  | N=12,247 | N=11,862 | N=385 |  |
| Physician age group, years |  |  |  |  |
| ≤ 44 | 4,320 (35.3%) | 4,188 (35.3%) | 132 (34.3%) | <.001 |
| 45-64 | 5,746 (46.9%) | 5,644 (47.6%) | 102 (26.5%) |  |
| 65-74 | 1,718 (14.0%) | 1,618 (13.6%) | 100 (26.0%) |  |
| 75+ | 454 (3.7%) | 404 (3.4%) | 50 (13.0%) |  |
| Missing | 9 (0.1%) | 8 (0.1%) | 1 (0.3%) |  |
| Physician age, years |  |  |  |  |
| Mean ± SD | 51 ± 13 | 51 ± 13 | 56 ± 16 | <.001 |
| Physician sex |  |  |  |  |
| Female | 5,861 (47.9%) | 5,674 (47.8%) | 187 (48.6%) | 0.775 |
| Male | 6,386 (52.1%) | 6,188 (52.2%) | 198 (51.4%) |  |
| Model of care |  |  |  |  |
| PEM: Non-team capitation | 3,114 (25.4%) | 3,052 (25.7%) | 62 (16.1%) | <.001 |
| PEM: Enhanced Fee-for-service | 2,854 (23.3%) | 2,769 (23.3%) | 85 (22.1%) |  |
| PEM: Team-based capitation | 2,893 (23.6%) | 2,825 (23.8%) | 68 (17.7%) |  |
| Non-PEM: Straight fee-for-service | 3,099 (25.3%) | 2,954 (24.9%) | 145 (37.7%) |  |
| Missing | 287 (2.3%) | 262 (2.2%) | 25 (6.5%) |  |
| Rurality (RIO) |  |  |  |  |
| Big cities 0 | 5,971 (48.8%) | 5,773 (48.7%) | 198 (51.4%) | 0.399 |
| Smaller cities (1 - 9) | 3,338 (27.3%) | 3,245 (27.4%) | 93 (24.2%) |  |
| Small towns (10 - 39) | 1,817 (14.8%) | 1,756 (14.8%) | 61 (15.8%) |  |
| Rural (40+) | 1,111 (9.1%) | 1,079 (9.1%) | 32 (8.3%) |  |
| Missing | 10 (0.1%) | 9 (0.1%) | 1 (0.3%) |  |
| Physician panel size group |  |  |  |  |
| <100 | 1,847 (15.1%) | 1,761 (14.8%) | 86 (22.3%) | <.001 |
| 100-499 | 1,378 (11.3%) | 1,310 (11.0%) | 68 (17.7%) |  |
| 500-999 | 2,369 (19.3%) | 2,287 (19.3%) | 82 (21.3%) |  |
| 1000-1499 | 2,980 (24.3%) | 2,917 (24.6%) | 63 (16.4%) |  |
| 1500-1999 | 1,899 (15.5%) | 1,858 (15.7%) | 41 (10.6%) |  |
| 2000+ | 1,487 (12.1%) | 1,467 (12.4%) | 20 (5.2%) |  |
| Missing | 287 (2.3%) | 262 (2.2%) | 25 (6.5%) |  |
| **Physician panel size** |  |  |  |  |
| Mean ± SD | 1,097 ± 829 | 1,106 ± 829 | 788 ± 741 | <.001 |
| Median (IQR) | 1,059 (417-1,580) | 1,069 (437-1,588) | 673 (114-1,259) | <.001 |
| **No. days with ≥ 1 billing** |  |  |  |  |
| March - Sept 2019 Mean ± SD | 100 ± 35 | 101 ± 35 | 73 ± 35 | <.001 |
| March - Sept 2020 Mean ± SD | 109 ± 42 | 109 ± 42 | 0 ± 0 |  |
| **No. Total visits** |  |  |  |  |
| March - Sept 2019 Mean ± SD | 2,061 ± 1,795 | 2,087 ± 1,803 | 1,266 ± 1,281 | <.001 |
| March - Sept 2020 Mean ± SD | 1,703 ± 1,674 | 1,758 ± 1,672 | 0 ± 0 | <.001 |
| **Percent of virtual visits** |  |  |  |  |
| March - Sept 2019 | 2.0 | 2.0 | 0.07 | <.001 |
| March - Sept 2020 | 66.0 | 66.0 | . |  |

**Notes:**
1. The total number of billing days for the study period including weekends and holidays was 203 days (29 weeks).

**Figure 1.**
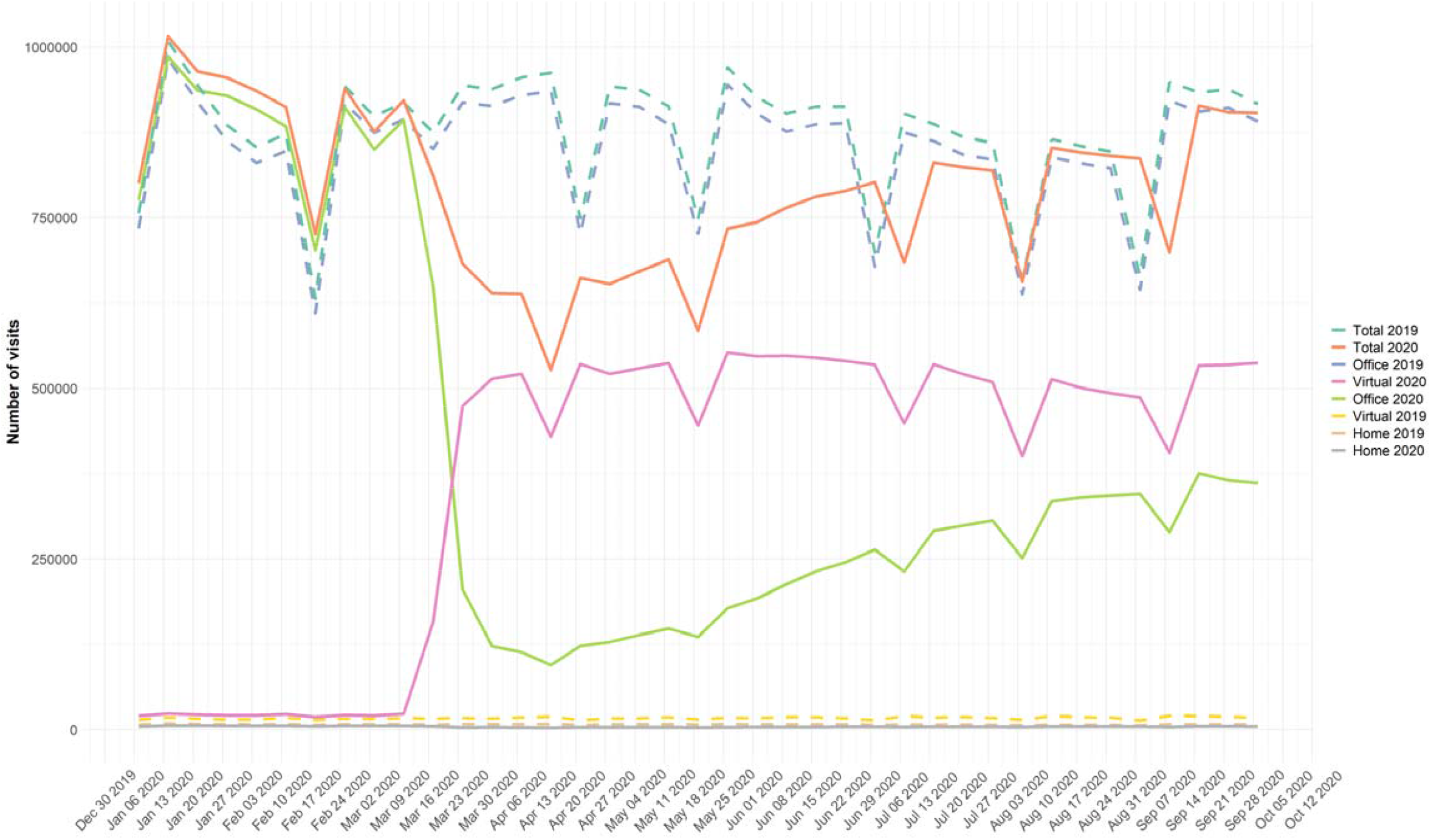
Primary care visits December 31, 2019 to September 29, 2020 compared to the same period in 2019 stratified by type of visit (office, virtual, home)

Figure 2 is a histogram of the number of physicians by ratio of total visit volume in the first 6 months of the pandemic to the total visit volume during the same period in 2019. The majority of family physicians had 0 to 50% fewer total visits post-pandemic compared to the same period in 2019.

**Figure 2.**
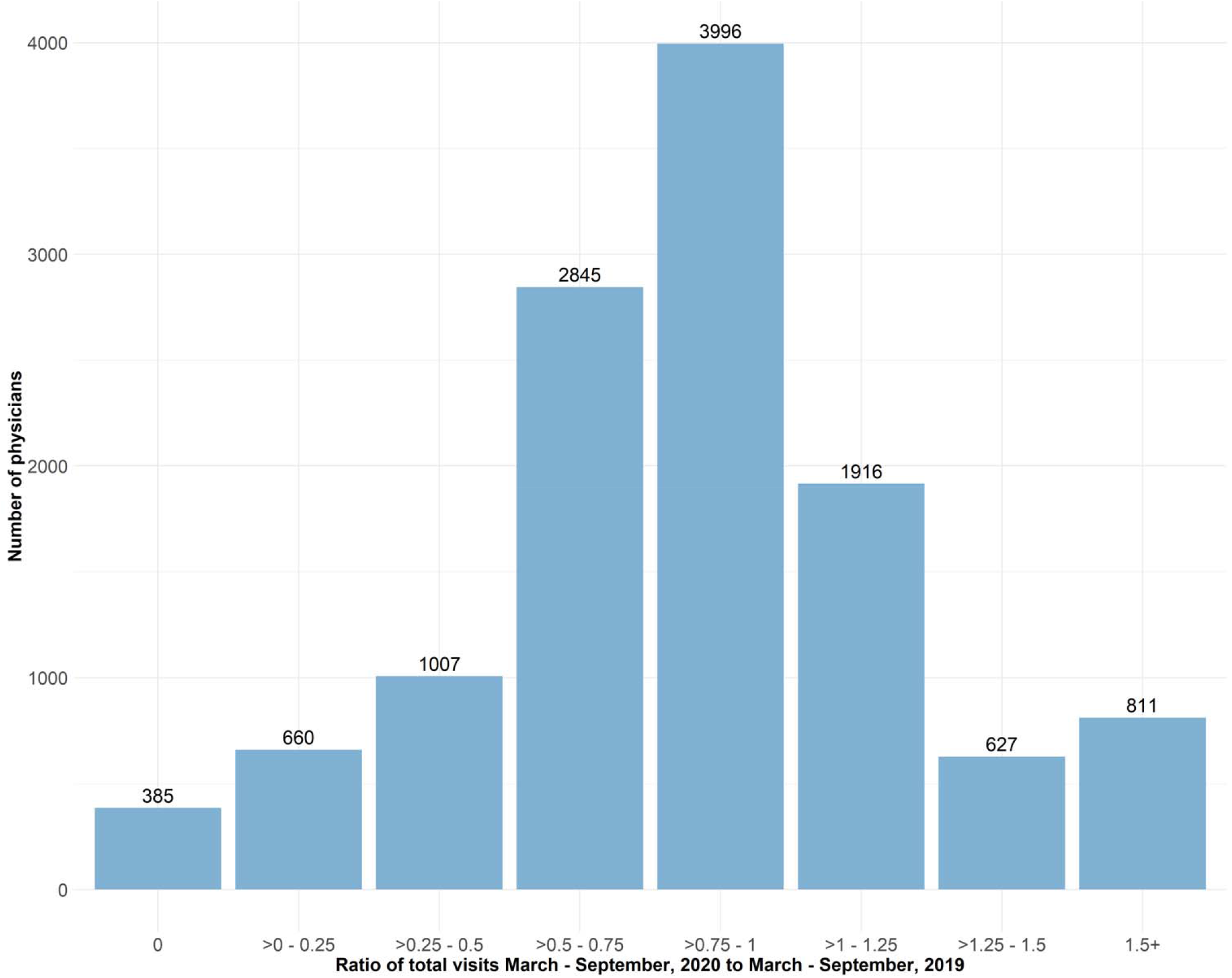
Histogram of the number of physicians by the ratio of total visits during the first six months of the COVID-19 pandemic (March to September 2020) to total visits during the same time period in 2019.

Figure 3 illustrates the large variation between and within patient enrolment models of the ratio of total visits during the first 6 months of the pandemic to the same period in 2019. The variation was not explained by model of care (ICC: 1.5%) or the specific practice group the physician belonged to (ICC: 4.6%)

**Figure 3.**
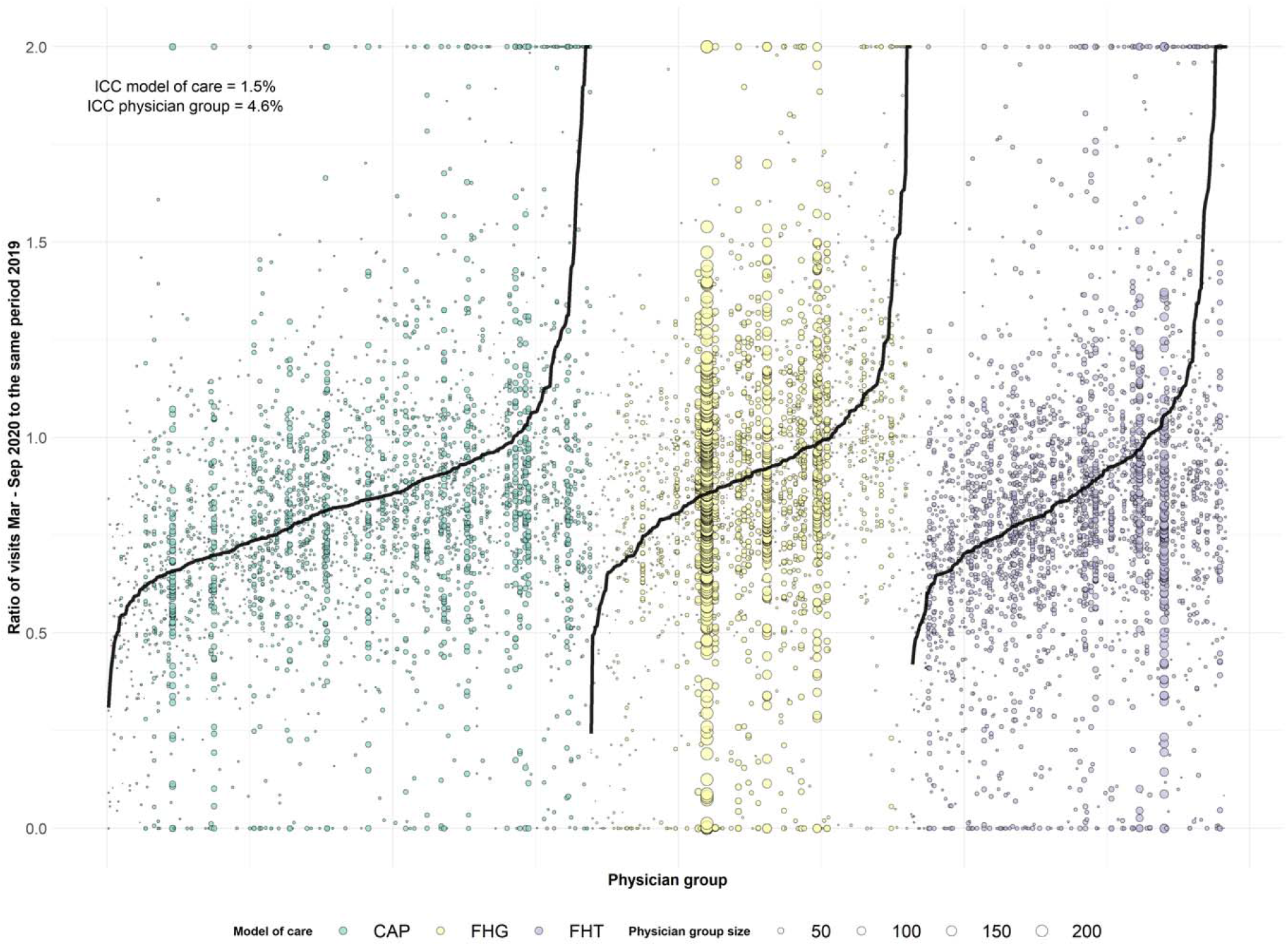
Variation within and between patient enrolment models in the ratio of total visits during the first six months of the COVID-19 pandemic (March to September 2020) to total visits during the same time period in 2019. Note: The black line represents the mean ratio for the practice group. Each group can have 3 or more physicians. Each dot represents a physician. Physicians within the same practice group are represented on the same vertical line.

There were 3.1% (n=385) of physicians practicing in 2019 that had no primary care visits during the first 6 months of the pandemic (Table 1). Compared with other family physicians, a higher portion of physicians with no primary care visits were age 75 or older (13.0% vs. 3.4%, p<0.001), practicing fee-for-service (38% vs 25%, p<0.001), had a panel size under 500 patients (40% vs 25%, p<0.001), had fewer billing days in 2019 (mean 73 vs. 101, p<0.001), and had fewer total visits in 2019 (1,266 vs 2,087, p<0.001). Fifty-six percent (n=215) of those who stopped working practiced in a patient enrolment model (PEM). The percentage of all family physicians who stopped work ranged from 0 to 14% in a given sub-region with higher percentages in both urban and rural areas (Figure 4).

**Figure 4.**
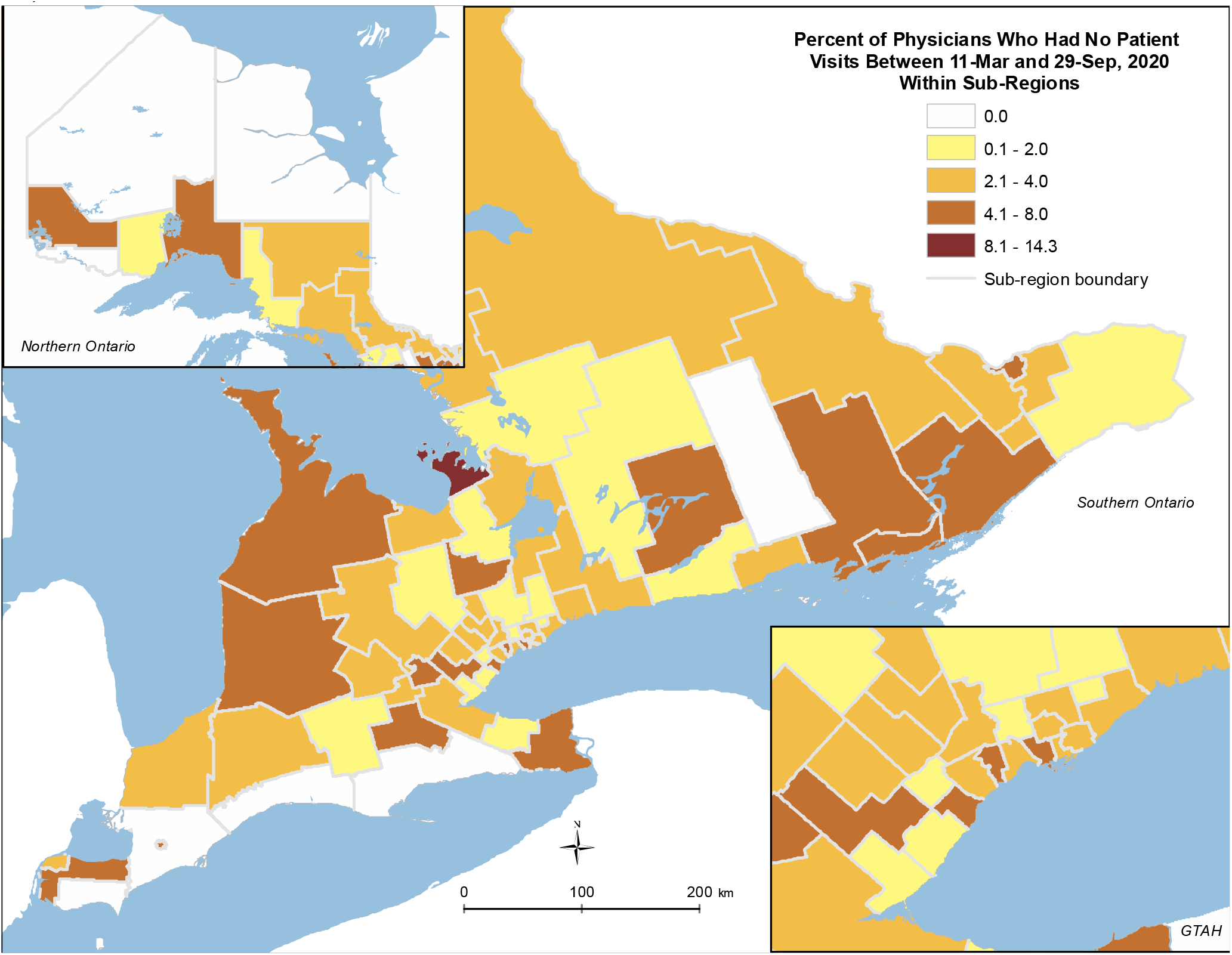
Percentage of all family physician within a sub-region who had no patient visits between March 11, 2020 and September 29, 2020

Figure 5 depicts the percentage of family physicians, between 2010 and 2020, who were practicing in January to March of the year but had no primary care visits between April to September of a given year. In the years from 2010 and 2019, an average of 1.6% of physicians stopped working entirely between April and September compared to 3.0% in 2020. The chi-squared test for independence was <0.001, rejecting the null hypothesis that the proportion of physicians stopping work is not dependent on the year.

**Figure 5.**
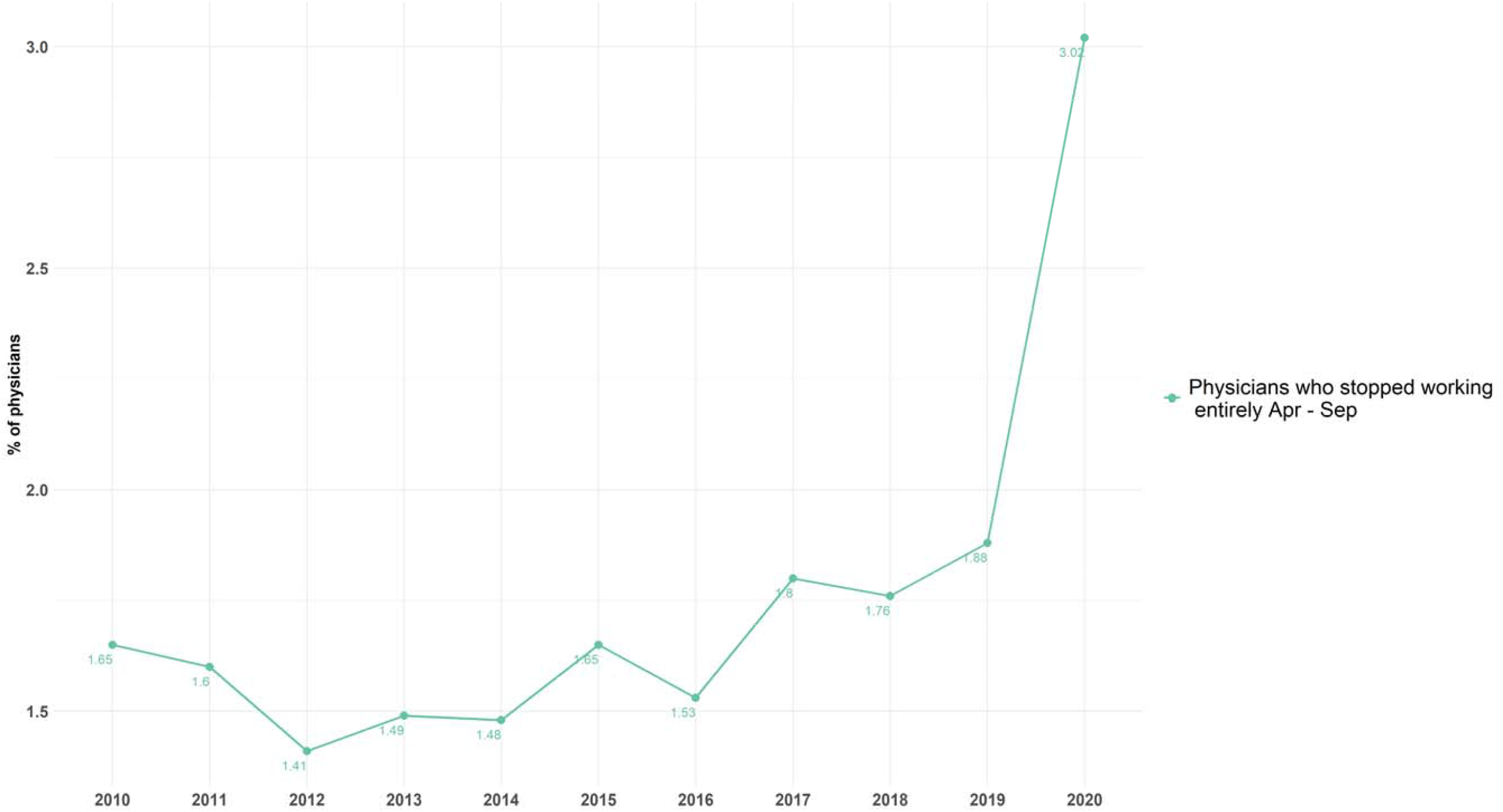
Percentage of family physicians who were actively practicing from January to March but who had zero primary care visits between April and September in the given year, 2010-2020

## Discussion

We conducted a population-based study assessing practice patterns of over 12,000 practicing family physicians in Ontario, Canada during the first six months of the COVID-19 pandemic. We found that roughly three percent stopped working during the first six months of the pandemic, approximately twice as many as in previous years. Our estimate of the proportion stopping work was consistent using two different analytic methods. Physicians stopping work were more likely to be over 75, practice fee-for-service, have a panel size under 500, and work less than other physicians in the previous year.

Although the absolute number of physicians stopping work was small, the impact on patients and communities is likely substantial. Just over half of the physicians who stopped working were practicing in a patient enrolment model and responsible for care of formally rostered patients. Using the mean panel size of 788, we can estimate these physicians cared for approximately 170,000 patients who may now be unattached. The number of unattached is likely even higher as some physicians who practiced outside a patient enrolment model and who stopped working were also providing comprehensive primary care. These health human resource challenges are occurring in the context of known doctor shortages in many urban and rural areas^16^ and where approximately 10% of the population do not have a family physician.^17^

We hypothesize that some family physicians accelerated their retirement plans because of the pandemic. Possible reasons include the concerns about health, increase practice costs due to recommended infection prevention and control measures, and drop in revenue due to the reduction in total visits. The drop in revenue would have been experienced most by those practicing in enhanced and straight fee-for-service models; not surprisingly, the pandemic has prompted physician leaders to redouble advocacy for payment reform.^18,19^ Burnout is another factor. Surveys done early in the pandemic reported that one-quarter of Canadian and half of American family physicians were exhausted.^5,8^ More recent surveys suggest almost one-third have persistent symptoms of burnout^20,21^—a signal that health human resource challenges will worsen.

We observed a large variation between physicians in the ratio of the number of visits in the first six months of the pandemic compared to the same period in the previous year. This variation was present between physicians practicing in different models, those in the same model but different groups, and even those within the same group practice. Our findings suggest that physicians largely operated independently when determining the degree to which they were able to continue providing care. This is likely multifactorial including considerations such as perceived personal risk for providing in person care, comfort with providing virtual care, variability in the interpretation of guidelines on the types of services that were considered essential, competing demands including delivery of COVID-19 related care in other settings and personal work-life balance issues related to the closures of schools, childcare and other areas of the economy impacting their families.

Our study has several limitations. First, we assessed the first six months following the pandemic and some portion of those who stopped working may have returned to work following that time. Even so, we still found higher levels of stopping work compared to the same period in previous years. Second, we did not distinguish between family physicians who practice comprehensive family medicine versus focused practice or walk-in medicine. However, it is likely that all those in patient enrolment models practice some portion of comprehensive family medicine.

## Conclusion

Approximately twice as many family physicians stopped work in Ontario, Canada during COVID-19 compared to previous years but the absolute number of physicians stopping work was small and those who stopped working had smaller patient panels. Our findings suggest COVID-19 may have accelerated retirement plans for a subset of older physicians with smaller, fee-for-service practices. More research is needed to understand the impact on primary care attachment and access to care.

## Data Availability

The data set from this study is held securely in coded form at the Institute for Clinical Evaluative Sciences (ICES). While data sharing agreements prohibit ICES from making the data set publicly available, access can be granted to those who meet pre-specified criteria for confidential access, available at www.ices.on.ca/DAS.

## Acknowledgements

Thank you to Peter Gozdyra for creating the map of percentage of physicians stopping work by subregion.

## Appendix. Fee codes used to virtually roster patients to primary care physicians

Patients were assigned to physicians first based on enrolment tables provided by the Ontario Ministry of Health. For patients who were not formally enrolled to a physician, we used virtual rostering. We assigned them to the physician who billed the highest amount for their care within the last two years based on the following primary care fee codes.

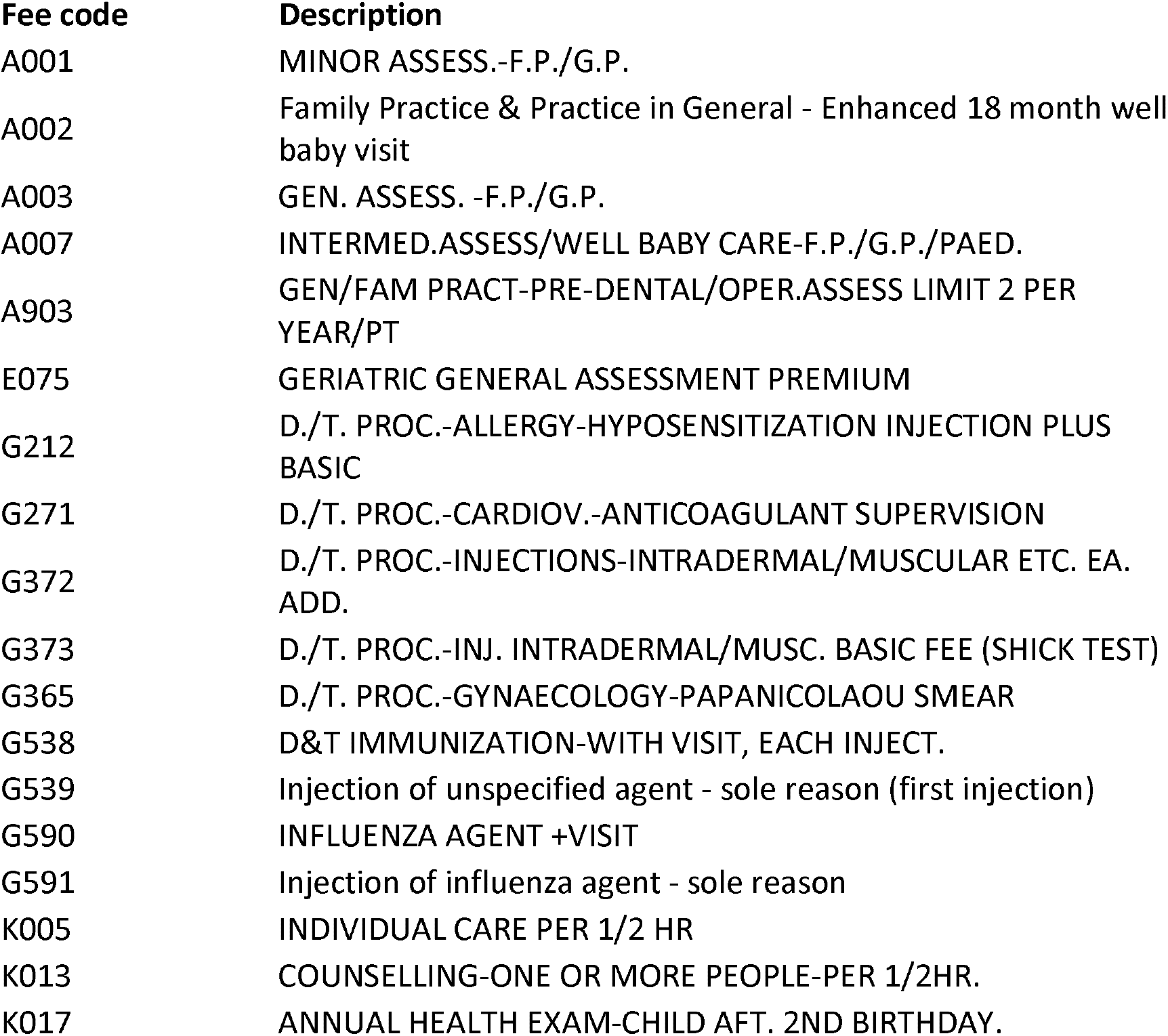
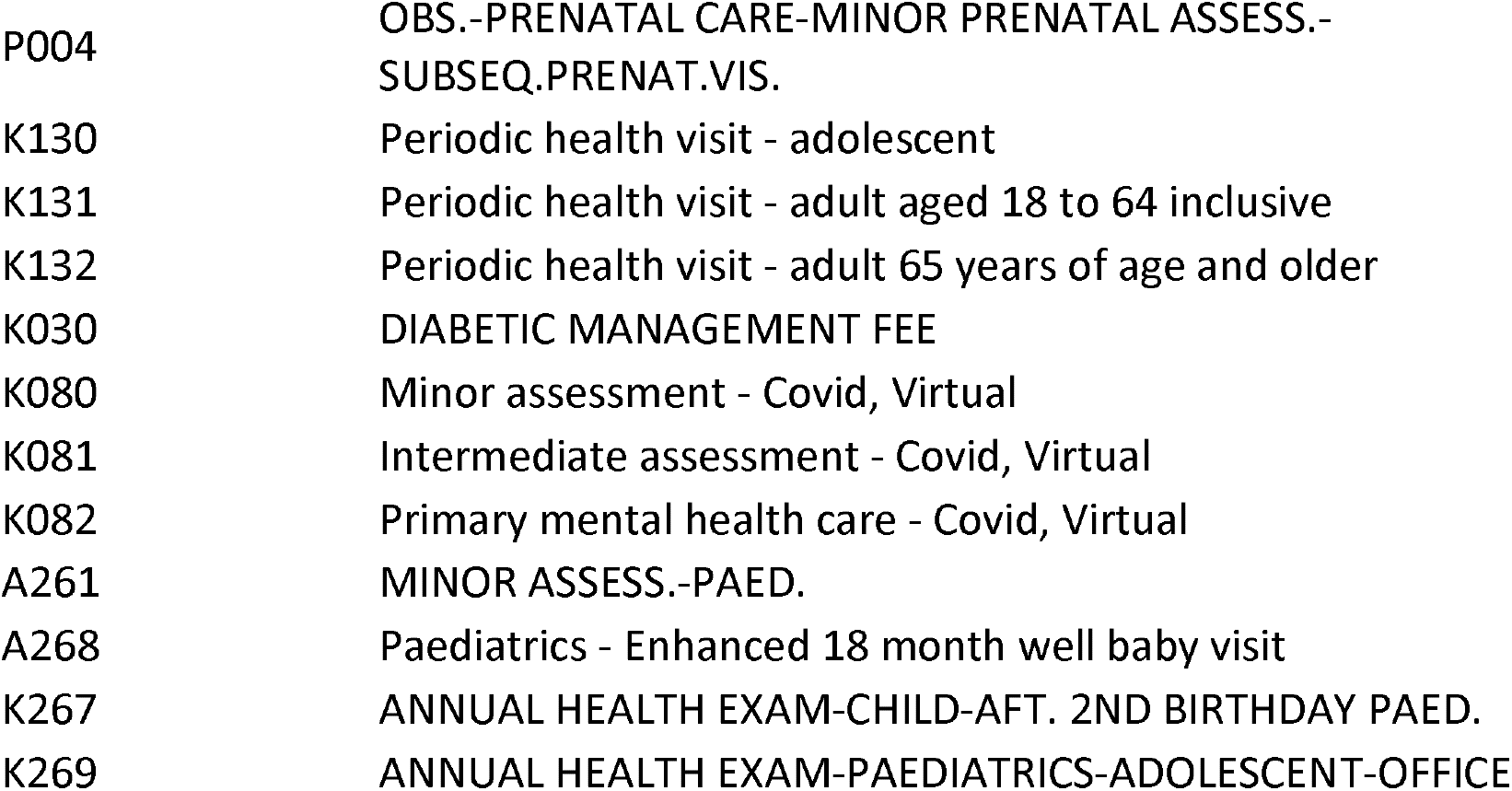

| Fee code | Description |
| --- | --- |
| A001 | MINOR ASSESS.-F.P./G.P. |
| A002 | Family Practice & Practice in General - Enhanced 18 month well baby visit |
| A003 | GEN. ASSESS. -F.P./G.P. |
| A007 | INTERMED.ASSESS/WELL BABY CARE-F.P./G.P./PAED. |
| A903 | GEN/FAM PRACT-PRE-DENTAL/OPER.ASSESS LIMIT 2 PER YEAR/PT |
| E075 | GERIATRIC GENERAL ASSESSMENT PREMIUM |
| G212 | D./T. PROC.-ALLERGY-HYPOSENSITIZATION INJECTION PLUS BASIC |
| G271 | D./T. PROC.-CARDIOV.-ANTICOAGULANT SUPERVISION |
| G372 | D./T. PROC.-INJECTIONS-INTRADERMAL/MUSCULAR ETC. EA. ADD. |
| G373 | D./T. PROC.-INJ. INTRADERMAL/MUSC. BASIC FEE (SHICK TEST) |
| G365 | D./T. PROC.-GYNAECOLOGY-PAPANICOLAOU SMEAR |
| G538 | D&T IMMUNIZATION-WITH VISIT, EACH INJECT. |
| G539 | Injection of unspecified agent - sole reason (first injection) |
| G590 | INFLUENZA AGENT +VISIT |
| G591 | Injection of influenza agent - sole reason |
| K005 | INDIVIDUAL CARE PER 1/2 HR |
| K013 | COUNSELLING-ONE OR MORE PEOPLE-PER 1/2HR. |
| K017 | ANNUAL HEALTH EXAM-CHILD AFT. 2ND BIRTHDAY. |
| P004 | OBS.-PRENATAL CARE-MINOR PRENATAL ASSESS.-<br>SUBSEQ.PRENAT.VIS. |
| K130 | Periodic health visit - adolescent |
| K131 | Periodic health visit - adult aged 18 to 64 inclusive |
| K132 | Periodic health visit - adult 65 years of age and older |
| K030 | DIABETIC MANAGEMENT FEE |
| K080 | Minor assessment - Covid, Virtual |
| K081 | Intermediate assessment - Covid, Virtual |
| K082 | Primary mental health care - Covid, Virtual |
| A261 | MINOR ASSESS.-PAED. |
| A268 | Paediatrics - Enhanced 18 month well baby visit |
| K267 | ANNUAL HEALTH EXAM-CHILD-AFT. 2ND BIRTHDAY PAED. |
| K269 | ANNUAL HEALTH EXAM-PAEDIATRICS-ADOLESCENT-OFFICE |

